# Robust metabolomic age prediction based on a wide selection of metabolites

**DOI:** 10.1101/2023.06.03.23290933

**Authors:** Tariq Faquih, Astrid van Hylckama Vlieg, Praveen Surendran, Adam S. Butterworth, Ruifang Li-Gao, Renée de Mutsert, Frits R. Rosendaal, Raymond Noordam, Diana van Heemst, Ko Willems van Dijk, Dennis O. Mook-Kanamori

## Abstract

Chronological age is a major risk factor for numerous diseases. However, chronological age does not capture the complex biological aging process. Biological aging can occur at a different pace in individuals of the same chronological age. Therefore, the difference between the chronological age and biologically driven aging could be more informative in reflecting health status. Metabolite levels are thought to reflect the integrated effects of both genetic and environmental factors on the rate of aging, and may thus provide a stronger signature for biological age than those previously developed using methylation and proteomics. Here, we set out to develop a metabolomic age prediction model by applying ridge regression and bootstrapping with 826 metabolites (of which 678 endogenous and 148 xenobiotics) measured by an untargeted high-performance liquid chromatography mass spectrometry platform (Metabolon) in 11,977 individuals (50.2% men) from the INTERVAL study (Cambridge, UK). Participants of the INTERVAL study are relatively healthy blood donors aged 18-75 years. After internal validation using bootstrapping, the models demonstrated high performance with an adjusted R^2^ of 0.82 using the endogenous metabolites only and an adjusted R^2^ of 0.83 when using the full set of 826 metabolites with age as outcome. The latter model performance could be indicative of xenobiotics predicting frailty. In summary, we developed robust models for predicting metabolomic age in a large relatively healthy population with a wide age range.

## Introduction

Chronological age is a major risk factor for a multitude of diseases (1, 2). The biology of aging comprises complex and multifactorial processes that are influenced by genetic, lifestyle and environmental factors (3-5). Evidently, the rate of biological aging varies between individuals, as in contrast to other individuals of the same age group, wherein, some individuals are able to live to an older chronological age without age-related diseases and disability (3). This suggests that chronological age does not align with the biological aging process. For this reason, several studies have aimed to capture the signature of biological changes due to aging process by predicting its rate based on biological factors using, for example, DNA methylation (6) and proteins (7).

Metabolomic profiling aims to identify small molecules that are mostly substrates or products of metabolism (metabolites). The number of metabolomics studies has increased in recent years due to major technological advances and the availability of (validated and high throughput) commercial and noncommercial analyses platforms. In addition, the current platforms have improved their capability to detect and quantify large numbers of endogenous and xenobiotic metabolites(8, 9). Since individual metabolomic profiles reflect the influences of both genetic and acquired factors, they are thought to provide a more holistic representation of biological processes, such as aging (8, 9). Furthermore, as metabolomic profiles are strongly affected by chronological age (4, 10) and sex (11, 12), they have been used to develop prediction models of chronological age (i.e., the metabolomic age) (13-16). However, predicting metabolomic age using metabolomics had limited success or faced methodological limitations due to several reasons. First, some studies used a relatively small sample size to predict age using hundreds or even thousands of metabolite predictors (10, 17). The inclusion of a larger number of predictors than the number of samples may cause overfitting and bias. Second, previous metabolomic age studies used targeted platforms, such as the Nightingale platform, that specifically measure a small number of lipoproteins and some low-molecular metabolites. This limits the variety of the metabolites and the information used for the prediction of age. Third, studies may be limited by the age distribution of the cohort study. This may restrict the model to a specific age range which affects the generalizability of the model in other studies and other age groups (10). Fourth, generalizability may also be reduced if the model is developed in cohorts with an oversampling of individuals with specific disease outcomes or specific population characteristics (7, 13). Fifth, statistical methodology such as stepwise selection, have been reported to cause overfitting of the prediction model (18). Finally, it is challenging to examine the model’s validity and generalizability in external studies or different populations particularly due to the dynamic nature of metabolite levels (19-21).

In this study, we aimed to develop a model to predict metabolomic age in a large healthy population with a widespread age range, using a single untargeted metabolomics platform. To attain this goal, we developed a prediction model using data from the INTERVAL study (22) (University of Cambridge, Cambridge, UK). Metabolomic profiles were available in 11,977 participants as measured using Metabolon’s (Durham, North Carolina, USA) untargeted metabolomics platform. These measurements included a broad range of endogenous and xenobiotic metabolites (n=1,363) from various biochemical pathways, thereby enabling the capture of metabolites related to a vast range of ageing effects.

## Methods

### INTERVAL Study

The INTERVAL study is a prospective cohort study of approximately 50,000 participants nested within a pragmatic randomized sample of blood donors (22). Between 2012 and 2014, blood donors, aged 18 years and older, were consented and recruited from 25 National Health Service Blood and Transplant (NHSBT) static donor centers across the UK. The INTERVAL study was approved by The National Research Ethics Service (11/EE/0538). Individuals with major disease (myocardial infarction, stroke, cancer etc.) as well as those who reported being unwell or having had recent illness or infection or did not fulfill the other criteria required for blood donation (22, 23) were ineligible for the study. Therefore, participants included in the study were predominantly healthy. Participants completed online questionnaires addressing basic lifestyle and health-related information, including self-reported height and weight, ethnicity, smoking status, alcohol consumption, doctor-diagnosed anemia, use of medications (hormone replacement therapy, iron supplements) and menopausal status (22). Untargeted metabolomic data were available in 11,979 individuals (age range 18 -75). Two individuals had incorrect or missing height/weight values and were therefore excluded from the study. Thus, the final sample size for the current study was 11,977.

### Untargeted metabolomic measurements

Untargeted metabolomic measurements were quantified at Metabolon Inc. (Durham, North Carolina, USA) using Metabolon™ Discovery HD4 platform. In brief, this process involves four independent ultra-high-performance liquid chromatography mass spectrometry (UHPLC-MS/MS) platforms (24, 25). Of these platforms, two used positive ionization reverse phase chromatography, one used negative ionization reverse phase chromatography, and one used hydrophilic interaction liquid chromatography negative ionization (25). Known metabolites were annotated at Metabolon Inc. with chemical names, super pathways, sub pathways, biochemical properties, and compound identifiers from various metabolite databases. Metabolomic measurements in the INTERVAL study were conducted in three batches (n=4087, 4566, and 3326). Subsequent harmonization and quality checks were performed between the batches by Metabolon. All metabolite measurements were scaled to a median of 1.

### Selection of Predictors

We aimed to select metabolites consistently and reliably measured by the Metabolon platform. In total 1,411 metabolites were measured in the INTERVAL study. First, we removed metabolites completely missing in at least one of the batches (n=175). Second, we excluded metabolites without annotation (unnamed metabolites) (n=258), keeping only endogenous and xenobiotic metabolites. These metabolites were excluded as they are inconsistently measured by the platform and are highly variable between batches and studies. Moreover, the lack of full annotation increases the uncertainty that we are using the same metabolite across batches and studies and leaves no secondary information for further verification. Third, we excluded metabolites measured in less than 100 individuals (n=30). Fourth, to ensure that the metabolites that are used for our model are not unique for the INTERVAL study, we cross-referenced our dataset with an independent study with Metabolon measurements. For this we used the Netherlands Epidemiology of Obesity (NEO) study, a population-based (n=599) cohort study of individuals aged 45–65 years (26, 27). Based on this comparison, we additionally excluded 122 metabolites that were not detected in the NEO study. The final set of metabolites included 826 metabolites, with 678 endogenous and 148 xenobiotic metabolites that were confirmed to be regularly measured by the Metabolon platform (Figure 1).

**Figure 1:**
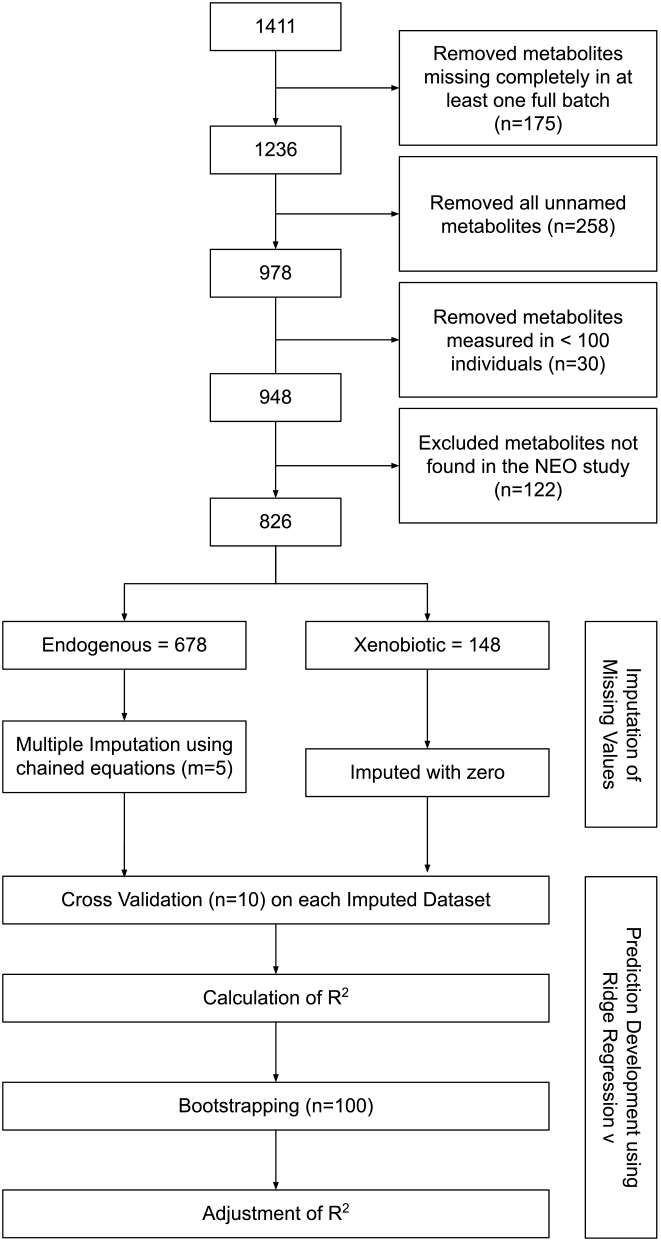
Flowchart of the selection of predictor metabolites and the development steps for the metabolomic age prediction model in the INTERVAL study

### Missing value imputation

Missing values were imputed using the pipeline as described in our previous work (27). In brief, endogenous metabolites were imputed by multiple imputation using chained equations method to generate five imputed datasets (m=5). For each metabolite with missing values, we used the outcome variable (i.e., age), 5-10 highly correlated related metabolites, body mass index (BMI), center number where the blood samples were collected, and the batch number to impute the missing values. Xenobiotic metabolites were imputed to zero to account for true missingness.

### Model Development

For the development of the model, we used ridge regression (28) to reduce potential overfitting. Cross validation was performed (n=10) in each imputed dataset to calculate the optimal shrinkage term (lambda). Subsequently the mean of the lambda values was used to develop the model on the stacked imputed datasets (i.e., all 5 datasets combined as one). Accordingly, the weight of the observations in the stacked dataset was set to 1/m = 1/5 = 0.2 (20). As the outcome (age) is a continuous variable, we assessed the fit of the model by deriving the R^2^. As an additional sensitivity test, we used generalized additive model (GAM) to examine and calculate the R^2^ for the nonlinear correlation. Internal validation was performed using bootstrapping (b=100). Bootstrapping results were used for the optimization of R^2^ and the calculation of the mean squared error (MSE) and the mean absolute error (MAE) of the model. Two models were developed using two sets of the selected metabolite predictors. First, we used the full set of endogenous and xenobiotic metabolites (n=826) to develop “model A”. Second, we used only the endogenous metabolites (n=678) to develop “model B”. As previous studies found that metabolomic profiles (12, 29) and aging (30-32) are influenced by the sex of individuals, we included sex as an additional predictor in both models.

### Sample size considerations

The primary database used to create the prediction model was the INTERVAL study (n= 11,977) with 826 predictors. We used the formulas described by Riley et al. (33) to confirm that our sample size (n) and number of predictor (p) are sufficient to minimize overfitting and provide high prediction precision. First, we calculated Copas global shrinkage factor for our data (34, 35) to see if it is above the recommended 0.9 threshold (33). Based on this calculation the estimated shrinkage factor was 0.95 if the adjusted R^2^ of the model was assumed to be 0.7. Second, we calculated the sample size required to ensure a small difference between the R^2^ and the adjusted R^2^ for the development model. Assuming the adjusted R^2^ was 0.7 again and a small desired R^2^ difference (R^2^_diff_=0.025), then the sample size required to achieve this should be at least n=9913. Third, we checked the sample size required for precise residual standard deviation of the model. Accordingly, we found the multiplicative margin of error (MMOE) to be less than 10% (MMOE =1.3%) using our n and p in the INTERVAL study. Finally, we checked the precision of the mean predicted outcome value (predicted age) of the model. We used n and p for the INTERVAL study and assumed that predicted age would have a mean of 45 and a variance of 35. Accordingly, the upper and lower bounds were approximately 45.34 and 44.65 respectively. Thus, the MMOE for the mean predicted outcome was less 1% (MMOE= 45.34 /45 = 1.007 = 0.7%). Therefore, the sample size of the INTERVAL study was optimal to minimize overfitting, optimism, and provide a precise estimation of the residual standard deviation and mean predicted values.

## Results

### Population characteristics

Characteristics of the total INTERVAL study population and age subgroups are summarized in Table 1. In total 11,977 individuals were included and had a normal age distribution with a mean age of 45 years and a range of 18 – 75 years. The number of men and women was approximately equal (50.2% were men). BMI was largely in the recommended range of 18.5-24.9 kg/m^2^ (36) with a mean BMI of 22.8 kg/m^2^ and was similar in all age groups.

**Table 1.** Characteristics of the INTERVAL study population.

|  | Total | 18 to 25 | 25 to 35 | 35 to 45 | 45 to 55 | 55 to 65 | 65 to 75 |
| --- | --- | --- | --- | --- | --- | --- | --- |
| N | 11977 | 1178 | 2245 | 2152 | 2867 | 2602 | 811 |
| Age (years), mean (range) | 45 (18-75) |  |  |  |  |  |  |
| Men, n (%) | 6019 (50.2%) | 485 (41.2%) | 965 (42.3%) | 1042 (48.4%) | 1560 (54.4%) | 1480 (56.9%) | 546 (67.3%) |
| BMI (kg/m<sup>2</sup>), mean (SD) | 22.8 (4.2) | 21.3 (4.0) | 22.0 (4.2) | 23.3 (4.4) | 23.5 (4.3) | 23.0 (3.9) | 22.6 (3.5) |

### Metabolomic Age Prediction

Prediction models A (endogenous plus xenobiotic metabolites) and B (endogenous metabolites only) were developed in the INTERVAL study using ridge regression. The workflow including metabolite selection, missing value imputation, and analyses are summarized in Figure 1. Internal validation using bootstrapping (resampling n = 100) and optimization provided an R^2^ of 0.83 (MSE=31, MAE=4.4) for model A, and 0.82 (MSE=33.7, MAE=4.6) for model B. The R^2^ for the generalized additional model (GAM) were slightly higher for both models, 0.85 for model A and 0.84 for model B (Figure 2, Supplementary Table 1, Supplementary Figure 1). Full tables with the intercept, sex and metabolite coefficients for model A and B are provided in Supplementary Table 2. This table also contains the mean values for the metabolites from the INTERVAL study.

**Figure 2:**
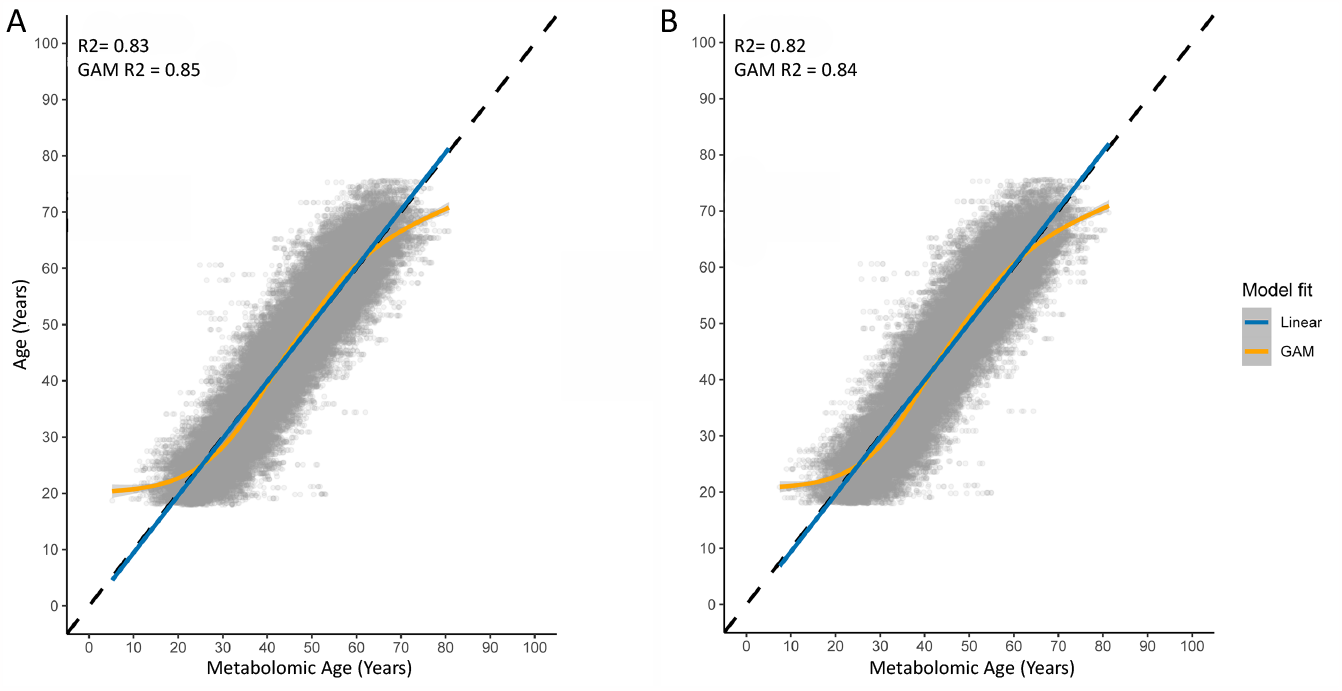
Correlation plots of the Metabolomic age (predicted age) on the horizontal axis and the chronological age on the vertical axis for model A (A) and model B (B). The data used is the stacked imputed datasets in the INTERVAL study. Abbreviations: GAM, generalized additive model.

## Discussion

In this study, we developed a prediction model for metabolomic age based on metabolite measurements, including a wide range of endogenous and xenobiotic metabolites belonging to a variety of biochemical pathways. The metabolomic measurements were performed in a single study using the same metabolomic platform and harmonized for within study and between batch variation. Importantly, we used multiple imputation, ridge regression, and bootstrapping (20) to develop and internally validate the metabolomic age prediction models. We developed two models, with the first model A using both endogenous and xenobiotic metabolites (n=826), and the second model B using the endogenous metabolites only (n=678). Both models had high adjusted R^2^ (model A = 0.83; Model B = 0.82), however, model B had slightly higher MSE and MAE, indicating higher error for the predicted values.

### Ridge Regression for Metabolomic Age

Our metabolomic age model is based on ridge regression which generates shrinkage factors that shrink metabolite coefficients with weak influence on the model to small values. Unlike methods such as LASSO and elastic net regression, this shrinkage never causes the coefficients to reach zero. Therefore, unlike those methods, ridge regression does not perform selection of predictors, in this case metabolites, due to the shrinkage term. Thus, the model consistently includes the same metabolites during the cross validation and internal validation by bootstrapping (20). Ridge regression thus ensures rigorousness and consideration of all the selected metabolite predictors. In addition, we found in our model that nonlinear GAM R^2^ to be slightly higher than linear R^2^ in both models. Figure 2 also shows that the GAM line curves at the edges of the age ranges. Therefore, predicting metabolomic age from this model in a nonlinear form, by adding a quadratic term or using a spline function, may further expand the model flexibility.

### Strengths and Limitations

A major strength of our current study is the development of robust metabolomic age prediction models based on a large number of metabolites measured in a large cohort with a wide age distribution. This sample size was confirmed to fit the required criteria for developing a model with low overfitting (33). Furthermore, the INTERVAL study included relatively healthy blood donors as participants. Blood donors are prescreened for a number of common diseases, making it a suitable study to develop a metabolomic age without being affected by specific disease-related effects on metabolites. Unlike previous metabolomic age predictors (alternatively referred to as metabolomic “clocks”), we developed our model using the latest Metabolon metabolomics platform that measures a large selection of metabolites. An advantage of this platform is the inclusion of xenobiotics, such as those derived from medication or pollution as well as endogenous metabolites. Thus, we were able to include metabolites originating from various internal and external sources. The xenobiotic metabolites represent some of the acquired environmental and lifestyle exposures of individuals that could play a role in biological aging. For example, medication metabolites could be a predictor of frailty. However, our study has two limitations. First, because the INTERVAL study does not have data for health-related outcomes, we could not assess the effects of different disease outcomes and health phenotypes on the metabolomic age. Second, as the INTERVAL study used here was cross sectional, we could not examine the model overtime or examine the ability of metabolomic age to predict disease outcomes or mortality in a future time point.

### External Validation for Metabolomic Age and Future Applications

Prediction models may suffer from overfitting due to selection bias, small sample size, methodology limitations, and lack of internal validation and calibration during development. However, another common issue regarding prediction modelling, such as the case with metabolomic age, is the challenge to externally validate them. This a common issue with prediction models in general. Indeed, few studies perform external validation of prediction models (19, 37). The reasons for the lack of external validation include the difficulty of applying and reproducing the prediction model method, lack of the full prediction variables to develop the model in the new dataset, or a lack of an appropriate sample size for external validation. Without external validation, the quality of the models cannot be properly assessed, and a model could still be overfitted despite presenting good results during internal validation(20, 21, 38). We took advantage of the sample size and wide age range to use a stringent ridge regression and internal validation method to develop the metabolomic age model. The resulting model demonstrated a high R^2^ for the metabolomic age. The high R^2^ from our models is unlikely to be due be overfitting as we considered and calculated the power and sensitivity of the model based on the sample size and number predictors (33), in addition to the rigorous prediction modelling methodology. The metabolomic age was able to predict chronological age well but not perfectly. The missing 0.2 from R^2^ is likely a reflection the biological aging process captured by the metabolomic age. However, future robust external validation, using weak and moderate calibrations in other populations (38) would be valuable for the metabolomic age models presented in this paper.

Several age prediction models have been developed that utilize different biological measurements such as targeted metabolomics (7, 14), proteomics (7), and epigenetics (DNA methylation)(6). These studies have reported that the metabolites were a strong predictor of body mass index, metabolic syndrome, cancer (7), type 2 diabetes, and cardiovascular disease (14). Furthermore, other biological clocks using proteomics and epigenetics were reported to be associated with depression and other outcomes (7). Therefore, it will be valuable to apply the prediction models presented in this paper in large longitudinal cohorts and examine the metabolomic age in relation to calendar age with the aforementioned phenotypes and other health related outcomes. Assessing the association between the residual between the chronological and metabolomic age with disease and health phenotypes overtime will provide clinically relevant prediction. The prediction strength will indicate the robustness of our model and provide the means to for external validation and re-calibration of the model.

In addition to our aim of addressing primary issues of the development of metabolomic age models, the Metabolon platform is expanded on the range of metabolites that can potentially be a better predictor of age. For example, previous metabolomic age studies have not measured or included xenobiotic metabolites. In our study, we were able to include this additional group of metabolites in model A. Furthermore, Model A may also be used in tandem with metabolomic age from targeted platforms, and biological clocks of different biological molecules measurements, similar to the work by Jansen et al. (7), to possibly improve or compare their predictive performance and their ability of capturing the effects of health-related phenotypes.

## Conclusions

We developed metabolomic age prediction models in a large relatively healthy population using a wide array of endogenous and xenobiotic metabolites. In model A with the endogenous and xenobiotic metabolites and in model B with endogenous metabolites only, the R^2^ of the linear fit was 0.82 and 0.83, respectively. We provided the full list of metabolites and their coefficients for both models. This data can enable other researchers to replicate our metabolomic age prediction model, externally validate it in their own studies with different disease outcomes and combine them with other age prediction models.

## Data Availability

Data is only available upon request.

## Acknowledgments and Disclosures

The authors of the NEO study thank all participants, all participating general practitioners for inviting eligible participants, all research nurses for data collection, and the NEO study group: Pat van Beelen, Petra Noordijk, and Ingeborg de Jonge for coordination, laboratory, and data management. The NEO study was approved by the medical ethical committee of the Leiden University Medical Centre (LUMC) and all participants provided written informed consent. The INTERVAL study was approved by The National Research Ethics Service (11/EE/0538) and all eligible donors provided a trial consent form before giving a blood donation.

## Author Contributions

T.O.F.- conceptualization, data curation, formal analysis, investigation, methodology, software, visualization, writing-original draft. R.L.-G.- validation, writing – review & editing. P.S. – supervision, conceptualization, project administration, resources, funding acquisition, writing – review & editing - A.S.B – project administration, resources, funding acquisition, writing – review & editing. R.d.M.- project administration, resources, funding acquisition, writing – review & editing. R.N. and D.V.H.- funding acquisition, writing – review & editing. F.R.R.- funding acquisition. A.v.H.V. and K.W.v.D- conceptualization, supervision, writing – review & editing. D.O.M.-K.- conceptualization, supervision, funding acquisition, writing – review & editing.

