## Supplementary figures and images for "Robust metabolomic age prediction based on a wide selection of metabolites"

### Supplementary Figure 1

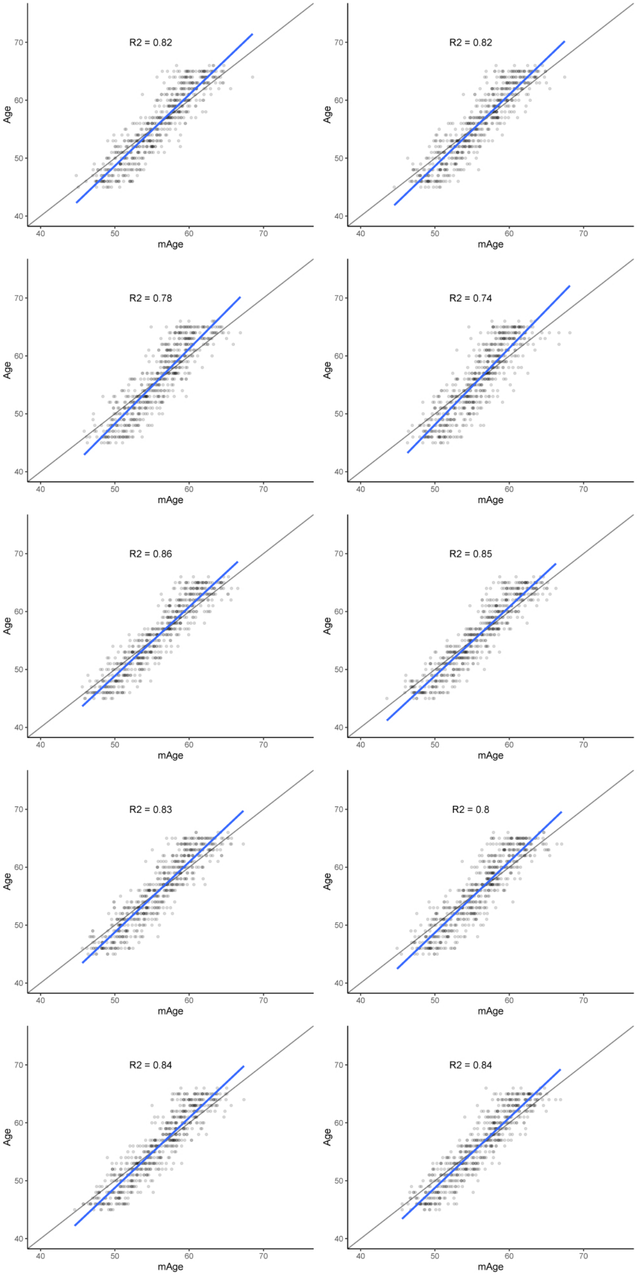
